# High-fidelity simulation versus case-based tutorial sessions for teaching pharmacology: convergent mixed methods research investigating undergraduate medical students’ performance and perception

**DOI:** 10.1101/2024.04.10.24305646

**Authors:** Rachid Kaddoura, Hanan Faraji, Farah Otaki, Rajan Radhakrishnan, Adrian Stanley, Agnes Paulus, Lisa Jackson, Reem Al Jayyousi, Sharon Mascarenhas, Meghana Sudhir, Jalal Alfroukh, Hardik Ghelani, Aida Joseph Azar, Amar Hassan Khamis, Reem Jan

## Abstract

**Introduction:** Medical educators strive to improve their curricula to enhance the student learning experience. The use of high-fidelity simulation within basic and clinical medical science subjects has been one of these initiatives. However, there is paucity of evidence on using simulation for teaching pharmacology, and the effectiveness of this teaching modality, relative to more traditional ones, have not been sufficiently investigated. Accordingly, this study compares the effects of high-fidelity simulation, which is designed in alignment with adult and experiential learning theories, and traditional case-based tutorial sessions on the performance and perception of undergraduate Year 2 medical students in pharmacology in Dubai, United Arab Emirates.

**Methods:** This study employed a convergent mixed methods approach. Forty-nine medical students were randomly assigned to one of two groups during the 16-week pharmacology course. Each group underwent one session delivered via high-fidelity simulation and another via a case-based tutorial. A short multiple-choice question quiz was administered twice (immediately upon completion of the respective sessions and 5 weeks afterwards) to assess knowledge retention. Furthermore, to explore the students’ perceptions regarding the two modes of learning delivery (independently and in relation to each other), an evaluation survey was administered following the delivery of each session. Thereafter, the iterative joint display analysis was used to develop a holistic understanding of the effect of high-fidelity simulation in comparison to traditional case-based tutorial sessions on pharmacology learning in the context of the study.

**Results:** There was no statistically significant difference in students’ knowledge retention between high-fidelity simulation and case-based tutorial sessions. Yet, students expressed a greater preference for high-fidelity simulation, describing the corresponding sessions as more varied, better at reinforcing learning, and closer to reality. As such, the meta-inferences led to expansion of the overall understanding around students’ satisfaction, to both confirmation and expansion of the systemic viewpoint around students’ preferences, and lastly to refinement in relation to the perspective around retained knowledge.

**Conclusion:** High-fidelity simulation was found to be as effective as case-based tutorial sessions in terms of students’ retention of knowledge. Nonetheless, students demonstrated a greater preference for high-fidelity simulation. The study advocates caution in adapting high-fidelity simulation, where careful appraisal can lend itself to identifying contexts where it is most effective.

## Introduction

In the ever-evolving landscape of medical education, schools strive to identify methods to enhance student learning and knowledge retention. Developing in health professionals a solid foundational understanding of the basic medical sciences is a given, and pharmacology is considered one of the core subjects. Learning pharmacology and therapeutics concepts has long posed challenges in medical education due to the complexity of drug-related information, including but not limited to core pharmacokinetic and pharmacodynamic principles, and therapeutics’ related information such as adverse reactions, and drug interactions and contraindications. Consequently, many medical students express difficulty in acquiring and retaining this knowledge (1). Hence, effective delivery of pharmacological concepts is crucial in preparing medical students to become safe prescribers (2). Pharmacology has been taught using various methods; the most implemented is traditional didactic lecture-based learning, which is characterized by a teacher-centric approach with minimal participation from and interaction with the students in the classroom. However, various studies have highlighted the challenges in establishing effective learning using this modality (3, 4). Therefore, different methods of teaching have been opted to enhance the student’s understanding and retention of pharmacology knowledge (4, 5). For example, problem-based methods such as case-based learning engage a small group of students in active, collaborative learning where they analyse cases or scenarios related to pharmacology that resemble real-world circumstances. This is usually done under the supervision of one or more tutor(s), while enabling, among the students, critical thinking and putting into practice the knowledge that they are acquiring (4, 6). Furthermore, laboratory practical sessions, which may involve using laboratory equipment or laboratory animals such as rodents, can provide direct experience in drug experimentation. This method allows the students to understand practical aspects of mechanisms of drug actions and pharmacological effects in a controlled setting (7).

Simulation-based medical education has been increasingly employed within medical curricula in recent years. It refers to any educational endeavour that employs simulation tools to recreate clinical situations. It allows for experimental learning and practice in a safe and controlled environment, without real-world consequences (8). Such technology serves as an additional resource in the students’ teaching, enhancing genuine encounters through controlled situations which trigger or imitate significant elements of reality (9). Furthermore, simulation emerges as a reliable tool for improving education, and facilitating uniform training and assessment in a safe environment, especially when effectively anchored in adult and experiential learning theories. These theories lie in the premise that adults deploy self-regulated learning (10, 11), are intrinsically motivated to learn, have mental models developed from previous experiences that form an increasing resource for learning, and regularly exercise analogical reasoning in learning and practice (12). This brings forward the Kolb’s experiential learning cycle (13, 14) which suggests that learning which occurs through a concrete and hands-on experience in a safe environment is followed by reflective observation (where the learner identifies gaps in their mental models). Next, the learner adapts their mental models (i.e., abstract conceptualisation), then actively experiments using the adapted mental models in a new experience. From this constructivist perspective (15), simulation provides a valuable resource for active experimentation. This cements new knowledge and long-term changes in practice. Among the previously identified limitations of the Kolb’s experiential learning theory is that it does not capture the learning that occurs in relating to others. The resulting simplistic view of experiential learning pulls it away from its origins, where it stemmed from human relations’ training (16). The literature emphasizes conceptualizing experiential education in more sociological terms, illustrating how the individual learner is inevitably connected to social, cultural, and/or environmental factors (17). Hence, from a practical perspective, it is worth deploying Kolb’s experiential learning theory in conjunction with a social constructionism theory where a small group of people learn through their social interactions. This emphasizes that participation and learning go together, and the learner is embedded in the context of learning (18).

While tutorial sessions are designed to affect the understanding of the theoretical background as well as practical skills in a highly interactive learning experience, simulation provides a dynamic platform where learners actively immerse themselves in lifelike scenarios, promoting practical application and critical thinking. Simulation offers a bridge between theory and practice, allowing learners to navigate complex situations, make decisions, and experience consequences in a controlled environment (19). Simulation also offers the opportunity for repeated practice and experimentation, while tutorials enhance theoretical knowledge by the traditional learning modalities. Learners can engage with simulation multiple times, refining their skills and strategies as they progress (20).

Whilst tutors aim to bridge the gap between theoretical knowledge and practical application, the choice between traditional classroom-based tutorial teaching and simulation-based learning emerges. Simulation-based learning may enhance the students’ understanding of the teaching materials and help in the practical part of medical education (21). However, it is still unclear, in certain disciplines, whether such high-tech teaching methods are sufficiently cost-effective. Furthermore, the incorporation of high-fidelity simulation into an already developed high-credit course such as pharmacology calls upon careful consideration of several factors. For instance, the integration of simulation teaching in the curriculum requires more time and resources from both faculty members and the institution, including higher costs compared to traditional methods (22). This is particularly relevant for high-fidelity simulation, which is a category of simulation that provides an exceptionally immersive learning experience. It uses advanced, usually interactive technology and realistic human mannequins, closely mimicking real-world scenarios (23, 24).

The current study uniquely integrates output of analysis around students’ academic performance (quantitative) with inferences made from analyses of data concerning their perception of two different teaching modalities (quantitative and qualitative). The fact that the respective high-fidelity simulation was designed in an evidence-driven manner, in alignment with experiential learning theories, further differentiates the current study. Lastly, the uniqueness of the study is further emphasized by the fact that it is conducted, among undergraduate Year 2 medical students, in a university of medicine and health sciences located in the Middle East and North Africa region (MENA). Hence, the overall purpose of the current study is to compare high-fidelity simulation to traditional case-based tutorial sessions for teaching pharmacology to undergraduate Year 2 medical students. This is done through investigating differences in knowledge retention of taught topics, and in students’ perception of the learning experiences and teaching modalities.

### Research questions

1. How did the learning experience of high-fidelity simulation and case-based tutorial sessions affect knowledge retention (i.e., performance) of pharmacology among Year 2 medical students?
2. How did Year 2 medical students perceive the learning experience of high-fidelity simulation and case-based tutorial sessions in the context of pharmacology teaching?
3. Which teaching modality (high-fidelity simulation or case-based tutorial sessions) is more effective in supporting Year 2 medical students’ learning of pharmacology?

## Methods

### Context of the study

This study was conducted at Mohammed Bin Rashid University of Medicine and Health Sciences (MBRU) in Dubai, United Arab Emirates, with a single cohort of Year 2 medical students. MBRU offers a six-year Bachelor of Medicine and Bachelor of Surgery degree (MBBS) that follows a spiral curriculum (25) which is divided into three phases: foundational basic sciences, preclinical sciences, and clinical rotations. Phase 1, which covers Year 1, serves as an introduction to fundamental medical concepts and basic human science. Phase 2, spanning Years 2 and 3, focuses on the different body organ systems in relation to clinical medicine. Years 4, 5, and 6 constitute Phase 3. During the first two years of Phase 3, students undertake clinical placements in different private and public hospitals. During Year 6, students undertake an apprenticeship, and assume greater clinical responsibilities under supervision (in preparation for residency). The study cohort was comprised of 49 Year 2 medical students in the academic year 2021-2022, aged 18-20 years (13 males and 36 females), enrolled in the ‘Principles of Pharmacology and Therapeutics’ course.

### Description of the intervention

The study revolved around the ‘Principles of Pharmacology and Therapeutics’ course which was delivered in the second semester (January - April 2022) of Year 2 of the MBBS program at MBRU. The corresponding semester ran over 16 calendar weeks, which included 3 teaching-free weeks [one for conducting in-course assessments, and two for the Spring break]. The weekly educational framework of this course consisted of two one-hour didactic lectures and one two-hour case-based tutorial. Case-based tutorial sessions were aimed to provide a more interactive learning experience, whereby students were divided into smaller sub-groups of 4-5 individuals, encouraged to address clinically-oriented cases and presented their work to the rest of the class, under the guidance and support of the pharmacology faculty members.

Two topics (which are usually taught through tutorial sessions) were selected by the research team for inclusion in the current study. These two topics were developed to be offered through two modalities: high-fidelity simulation and case-based tutorial sessions. The first topic was Routes of Administration (ROA), and its learning objectives were to (i) compare and contrast onset and duration of action of a drug administered through different routes, and (ii) identify the best route of administration for a patient based on patient-specific information. The second selected topic was Drug Toxicity and Interactions (TOX), and its learning objectives were to (i) identify drug interactions, recognize the symptoms of drug overdose, (ii) identify their consequences, (iii) suggest management strategies, and (iv) describe the mechanisms underlying them. Both sessions were adapted to high-fidelity simulation, using identical learning objectives and including three identical cases/scenarios, assuming the learners are self-regulated and intrinsically motivated to learn from experiences. To reinforce that, the learners were offered pre-session readings (e.g., treatment guidelines and drug information) to enable them to maximize the learning during the respective sessions. The design of those sessions was based on theoretic and empiric research in adult and experiential learning, where the educators took into account the learners, their experiences, and the overall learning environment (12). The high-fidelity simulation involved approximately eight members of staff, including three medical and/or pharmacology faculty members, three stimulation technicians, and two members of staff for ushering the student groups to their respective scenario rooms and for facilitating the end-of-session assessments and surveys. The entailed learning facilitation was in alignment with Kolb’s Experiential Learning Theory (14, 26). Each scenario required one medical or pharmacology faculty member to lead the scenario, and one simulation technician to operate the mannequin, or one standardized patient. The case-based tutorial session involved one or two pharmacology faculty member(s), who facilitated the learning during that session, assuming that the learners are self-regulated and are intrinsically motivated (12).

The first topic, ROA, was delivered during Week 4 (31^st^ January 2022). For this session, the students were split into two groups: the first group attended the high-fidelity simulation, and the second group attended the case-based tutorial session. The second topic, TOX, was delivered during Week 10 (14^th^ March 2022). For this session, the second group attended the high-fidelity simulation whereas the first group attended the case-based tutorial. For both high-fidelity simulation and case-based tutorial sessions, students were sub-divided into smaller groups of 4-5 individuals who rotated through the scenarios and worked on solving the corresponding cases together. The high-fidelity simulation sessions took place at the Khalaf Ahmad Al Habtoor Medical Simulation Center within MBRU. These sessions utilized high-fidelity mannequins and/or standardized patients that manifested physical examination signs, or could be questioned about symptoms, respectively. Additionally, the mannequins had continuous vital signs monitor displays which could be manipulated by the operator. The other more traditional teaching method was a case-based tutorial, which was delivered in a classroom setting, with no simulated patients present or any high-tech module involved. A brief timeline of the course delivery is illustrated in Figure 1.

**Figure 1.**
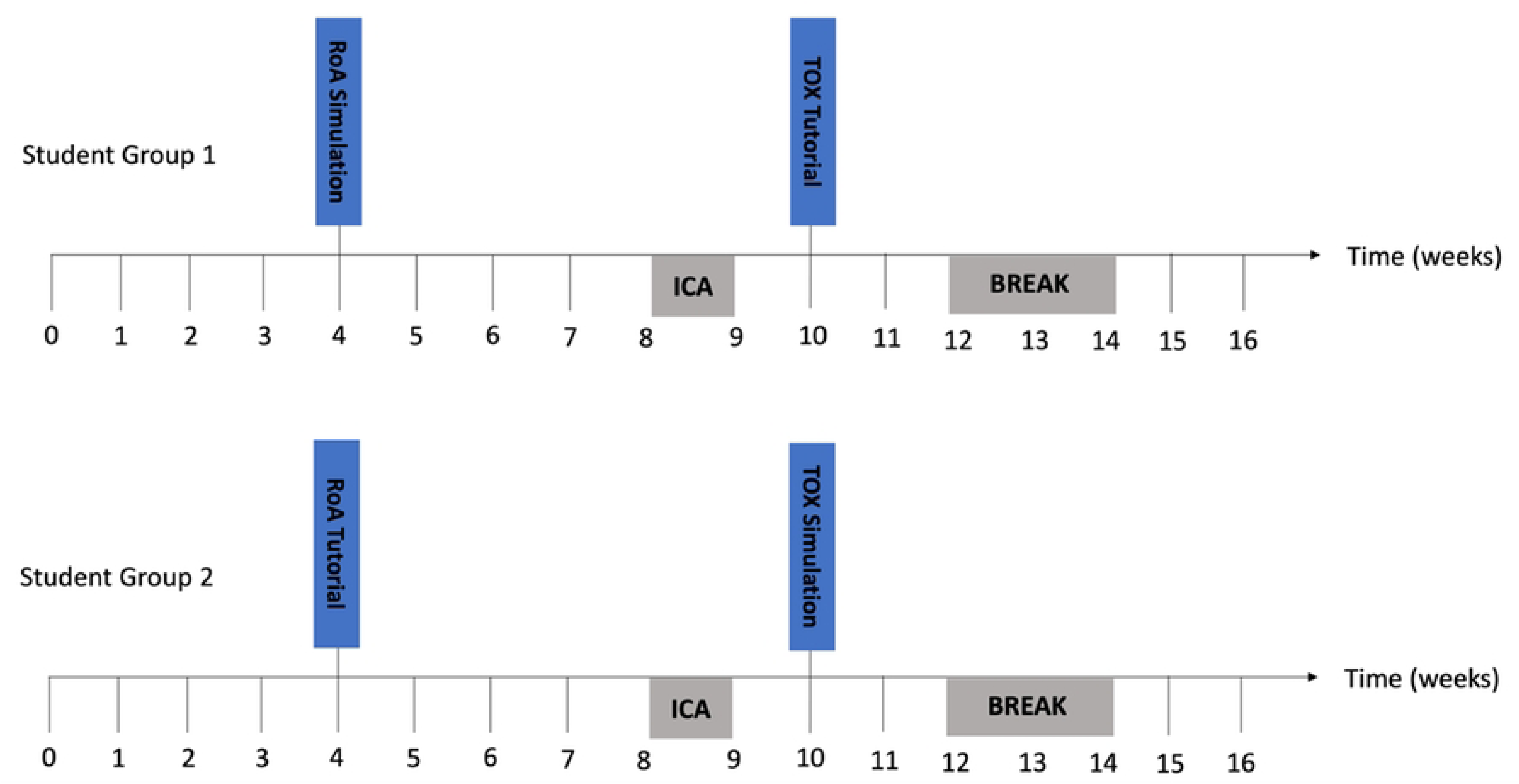
Timeline of the delivery of the two pharmacology topics: Routes of administration (ROA) and drug toxicity and interactions (TOX). ROA was delivered to Student Group 1 via high-fidelity simulation and to Student Group 2 via a case-based tutorial during the fourth week of the course. TOX was delivered during the tenth week of the course to Student Group 1 via a case-based tutorial and to Student Group 2 via high-fidelity simulation. ICA, In-course assessment.

### Research design

The current study relied on a convergent mixed methods research design (27, 28), which has been encouraged in the health professions’ education research field, and provides a more comprehensive view of the topic under investigation (29, 30). Therefore, three data sources were used to address the study’s research questions: (i) a 10 multiple-choice question (MCQ) quiz administered at two time points: a. At the end of each session (to assess short-term knowledge retention), and b. Five weeks after the session (to assess long-term knowledge retention) - quantitative data, (ii) a qualitative survey immediately after each session, and (iii) a quantitative survey after the students completed the two sessions (irrespective of the order) to capture their perception of the experience. Quantitative and qualitative data, from the three data sources, were analysed independently, and then the generated inferences were integrated to result in meta-inferences using the iterative joint display analysis process (26). Based on various knowledge transfer and translation frameworks, this study design taps into more than one level of analysis. For example, according to the Kirkpatrick evaluation model, this study’s research design captures Level One (i.e., reaction- perception) and Level Two (i.e., learning- knowledge retention/ performance) (31). In terms of the Learning-Transfer Evaluation Model (LTEM), the research design of the current study covers Tiers One through Four (namely: Attendance, Activity, Learner Perceptions, and Knowledge) (32). Accordingly, this data merging is believed to raise the reliability of the study, offering a systemic point of view of the subject matter (33). Ethical approval for the current study was granted by the Mohammed Bin Rashid University of Medicine and Health Sciences-Institutional Review Board (MBRU-IRB-2020-001), and electronic informed consent was obtained from all participants prior to participating in the research study.

### Data collection

A summarized descriptive overview of data collection over the study timeline is depicted in Figure 2, where all the data was collected 1^st^ January through 30^th^ April 2022 (around the time of the respective course delivery).

**Figure 2.**
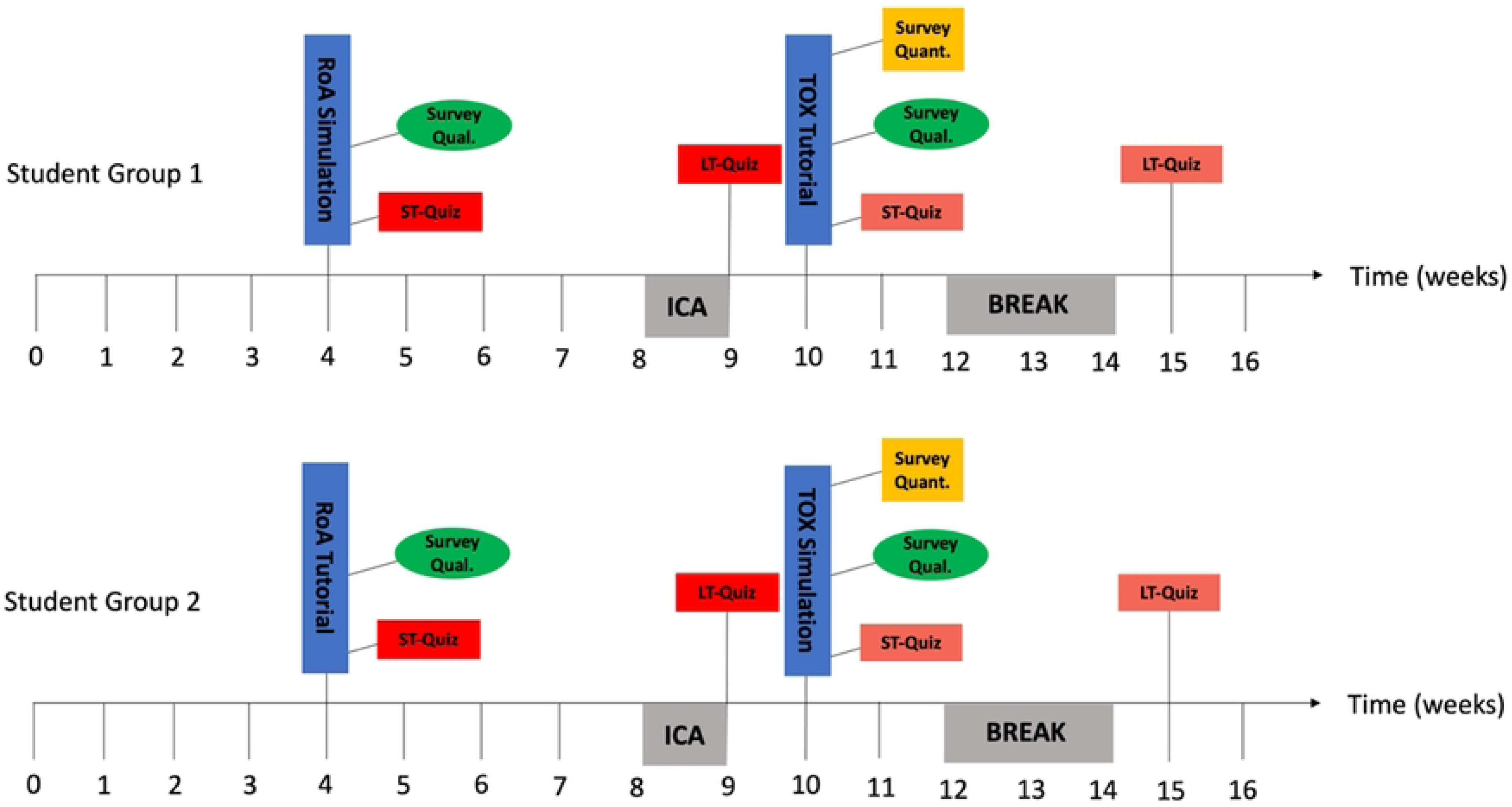
Data collection points superimposed on the timeline of the course delivery. ROA, Routes of administration topic; TOX, drug toxicity and interactions topic; ST-Quiz, Short-term quiz; LT-Quiz, Long-term quiz; ICA, in-course assessment; Survey Quant., quantitative survey; Survey Qual., qualitative survey.

#### Quantitative data source- students’ performance

To assess knowledge retention, a 10-MCQ quiz was used. The quiz was developed by two pharmacology faculty members, and reviewed by a panel of three pharmacology experts (two basic and one clinical) and two clinical faculty members in the fields of family medicine and internal medicine. Any modifications were addressed to ensure the accuracy and validity of the questions. The 10-MCQ quiz was delivered to the students, via an online Learning Management System (LMS), immediately after their assigned session (high-fidelity simulation or case-based tutorial) assessing knowledge learned while participating in the respective session (short-term knowledge retention). Unbeknownst to the students, the same 10 MCQ test was delivered again 5 weeks following the learning session to assess their long-term knowledge retention of the information learned. Students were not briefed about the long-term knowledge retention test to minimize bias related to any potential test preparation. In total, each student completed four quizzes, their scores were recorded and computed, awarding one point for each correct answer and zero points for each incorrect answer. The total score was calculated for each student as the sum of the correct answers, which (in the context of this study) is a reflection of the students’ performance/ knowledge retention.

#### Qualitative data source- students’ perception

To assess the students’ perception of both modalities, a descriptive survey was administered following the delivery of each session. The testimonies of students were collected to explore the students’ perceptions regarding the two teaching modalities. Each entry was assigned a unique identifier, composed of two parts. A serial number (i.e., 01 to 90), followed by ‘S’ for high-fidelity simulation or ‘T’ for case-based tutorial. For example, the identifier: 21S, represents the entry number 21, which corresponds to a student’s feedback after their experience with high-fidelity simulation. Because the data collection was anonymized, if the same student chose to provide feedback after their experience with case-based tutorial, their comment will be assigned another number, followed by ‘T’ for case-based tutorial.

#### Quantitative data source- students’ perception

The corresponding survey was administered immediately after the second topic (TOX sessions). This timing was chosen because, by that point, all students had experienced both the high-fidelity simulation and case-based tutorial session. The survey consisted of two independent dichotomous questions: (i) which is your preferred method of learning? and (ii) which method do you consider more useful?, where students were asked to choose between the two teaching modalities: high-fidelity simulation or case-based tutorial sessions.

### Data analysis

#### Quantitative data- students’ performance

This quantitative data was analysed using SPSS for Windows Version 27. The descriptive analysis consisted of computing the Change in Knowledge Retention (Short-term Knowledge Retention minus Long-term Knowledge Retention). Then, the mean and standard deviation for the Short-term Knowledge Retention and Long-term Knowledge Retention, along with the newly computed variable: Change in Knowledge Retention, were calculated. To select the appropriate inferential analysis tests, a test of normality was conducted for each of the three variables: Short-term Knowledge Retention, Long-term Knowledge Retention, and Change in Knowledge Retention. The data were all found to be not normally distributed. Accordingly, the nonparametric test of two independent samples: the Mann-Whitney U test, was used to assess the potentiality of associations between the following variables: the Short-term Knowledge Retention, Long-term Knowledge Retention, and Change in Knowledge Retention, and teaching modality (i.e., high-fidelity simulation or case-based tutorial sessions).

#### Qualitative data- students’ perception

The data analysis was started after the conclusion of the data collection phase. The dataset constituted a consolidation of all the entries from the qualitative surveys (conducted right after the respective sessions). The data was inductively analysed, by two researchers (H.F. and R.K.), employing a participant-centred, phenomenological approach to thematic analysis (34). The researchers proactively acknowledged potential factors influencing their perceptions of the subject matter. Consistency was rigorously maintained throughout the analysis process, adhering to an iterative approach rooted in constructivist epistemology (35). Unlike conventional scientific inquiry, this interpretative process necessitated acknowledging and reenacting the participants’ lived experiences. The focus of the chosen approach is not on establishing causal explanations, but on comprehending the participants, including their attitudes, behaviours, and actions (36). This methodology presupposes that participants’ thoughts can be apprehended by interpreting and, in turn, gaining a comprehensive understanding of their self-expression.

The process of analysis followed the six-step framework initially introduced by Braun and Clarke (37). This multi-phased approach to inductive qualitative analysis is encouraged in socio-behavioural research, in general, and health professions education research, in specific (38). NVivo software version 12.0 plus (QSR International Pty. Ltd., Chadstone, Australia) was utilized to code the data, and in turn, accelerate the classification of the identified text segments.

In the first phase of the analysis, the two researchers immersed themselves in the data. They jointly read through the deidentified comments, initiating a process of reflection on their content to properly familiarize themselves with the data. In the second stage, the transcripts were carefully examined. Text fragments pertaining, either directly or indirectly, to the research question were identified and extracted. Essentially, any segment of text that reflected on the students’ personal encounters with the modalities: high-fidelity simulation and/ or case-based tutorial sessions, as well as their involvement with the course material, was marked for reference and assigned a preliminary code. This continued until no further revelations emerged from the dataset, indicating that data saturation had been reached.

This process led to the classification of coded text fragments, paving the way for the researchers to embark on the third phase of analysis. This involved multiple rounds of reflection and exploration of potential connections, leading to the emergence of several potential constellations of categories. In the fourth stage, the researchers determined the most effective method for amalgamating the categories into overarching themes (Figure 3). All themes and categories were subsequently coded and defined in the context of the current study, marking the completion of stage five. The result of this process formed the study’s conceptual framework, providing guidance for the sixth and final phase of the thematic analysis: the narrative presentation of the findings, in accordance with established guidelines (39). To enhance the credibility of the findings, the researchers conducted a count and documented the number of text fragments within each category of the identified themes. If a single participant contributed more than one pertinent text fragment, all were consolidated into one entry. Essentially, the tally represents the number of participants who reflected on matters pertinent to the respective categories.

**Figure 3.**
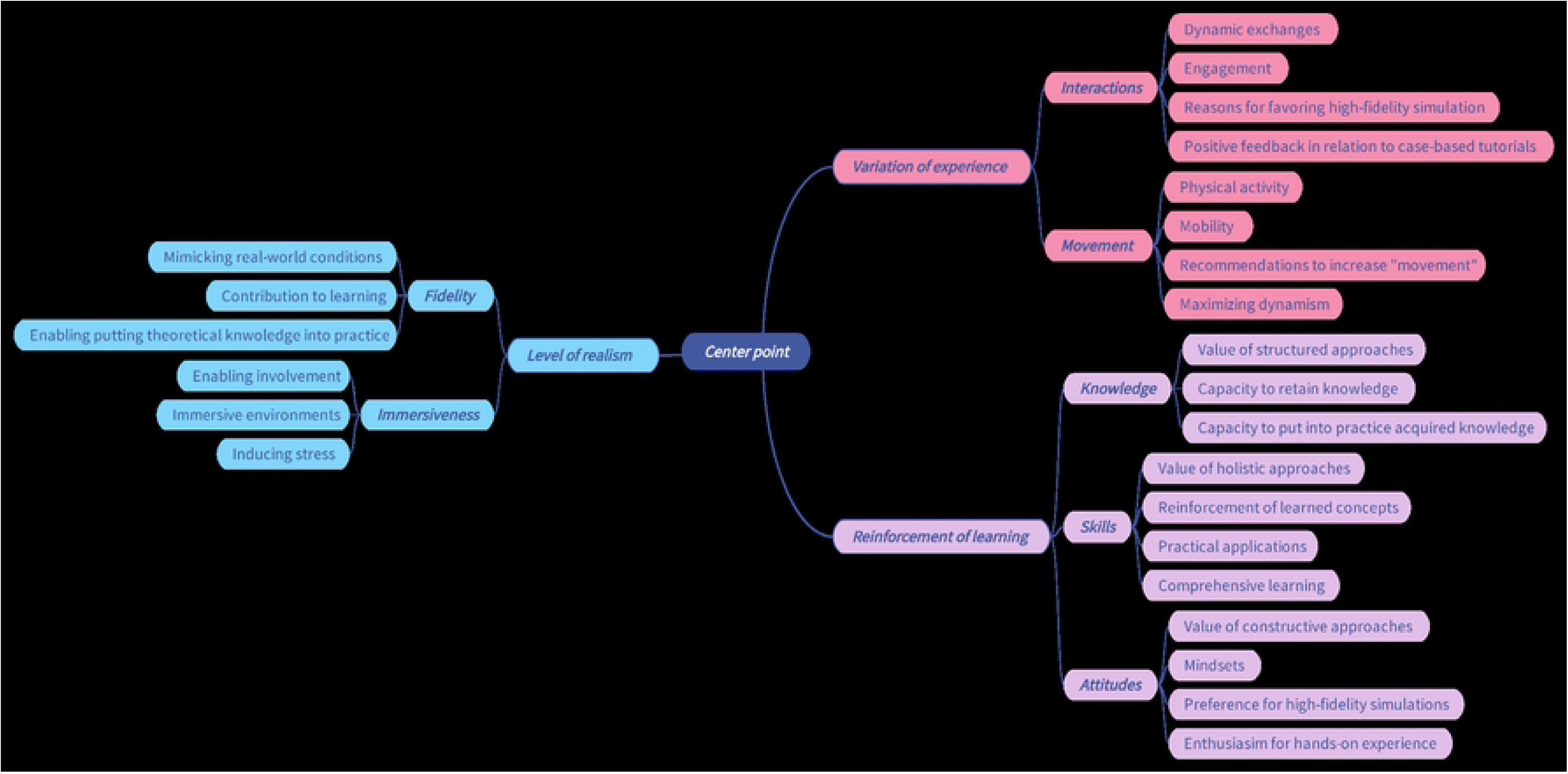
Mind map deployed as a tool to facilitate the qualitative analysis.

#### Quantitative data- students’ perception

This quantitative survey data was analysed descriptively using SPSS for Windows Version 27. The analysis consisted of computing the proportions for the two variables.

### Integration

Following the completion of the independent data analyses of quantitative and qualitative data, the generated findings were integrated using the iterative joint display analysis process (26). This stage allowed for drawing meta-inferences from the mapping of findings generated from each of the independent preceding analyses. In alignment with the guidelines of reporting on mixed methods research (39) that were adhered to for the current study, the output of the integration was meant to address the third research question inquiring about the effectiveness of the two modalities in relation to each other. Accordingly, within the context of the current study, effectiveness can be defined as the extent to which the respective learning and teaching interventions (i.e., high-fidelity simulation and case-based tutorial sessions) met their preset educational outcomes, taking into account the students’ performance and personal reflections on their lived experiences. The generation of meta-inferences enabled the researchers to identify where the findings build upon (or at least confirm) each other, as well as where they contradict each other.

## Results

In alignment with the guidelines of reporting on mixed methods research (39) that were adhered to for the current study, the analysis of data related to the students’ performance addressed the first research question of the current study, while the output of analysis of perception data answered the second research question. Furthermore, as previously mentioned in the Methodology section, the third research question was addressed through the integration of findings around students’ performance and perception.

### Output of quantitative analysis- students’ performance

Out of 49 learners, 43 learners experienced both teaching modalities (whereas 6 learners did not experience the case-based tutorial teaching modality). The short-term knowledge retention test was completed 92 times (by 49 learners post-simulation and 43 learners post-tutorial), while the long-term knowledge retention quiz was completed 73 times (by 35 learners post-simulation and 38 learners post-tutorial) (Table 1).

**Table 1.**
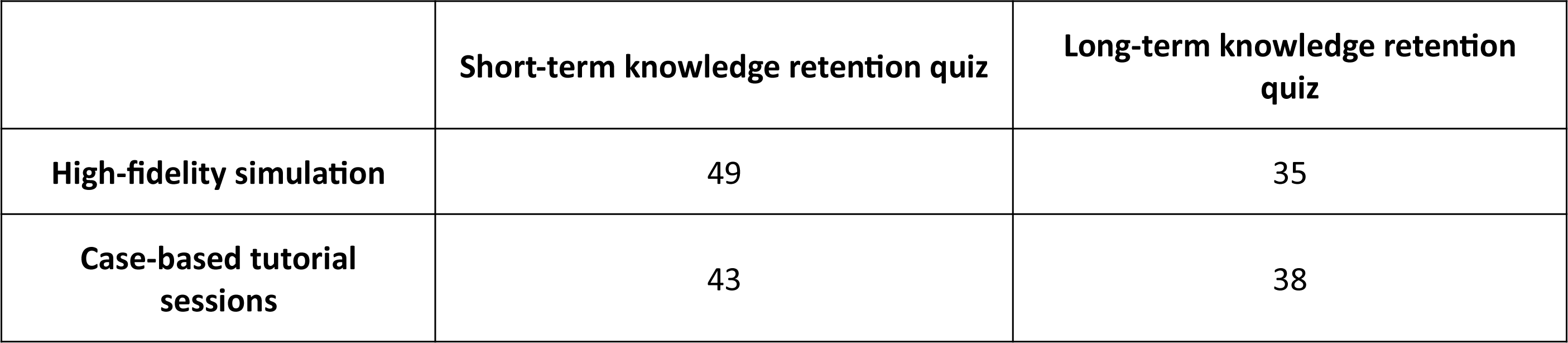
Distribution of learners across teaching modalities vis-à-vis quizzes.

There were no statistically significant differences in students’ short-term knowledge retention, long-term knowledge retention, and change in knowledge retention between teaching modalities (high-fidelity simulation or case-based tutorial sessions) (*p* > 0.05; Table 2).

**Table 2.** Short-term and long-term knowledge retention and change in knowledge retention in high-fidelity simulation versus case-based tutorial sessions.

| | Short-term Knowledge Retention score mean ± SD (%) ($p = 0.49$) | Long-term Knowledge Retention score mean ± SD (%) ($p = 0.59$) | Change in Knowledge Retention (ST-LT) score mean ± SD (%) ($p = 0.69$) |
| --- | --- | --- | --- |
| High-fidelity simulation | 78.79 ± 17.19 | 77.71 ± 23.77 | 2.57 ± 26.05 |
| Case-based tutorial sessions | 81.16 ± 15.62 | 77.37 ± 19.82 | 2.89 ± 17.99 |

### Output of qualitative analysis- students’ perception

The total number of entries that underwent the qualitative analysis were 90: 48 were collected from students immediately after their experience with high-fidelity simulation and 42 were collected from students immediately after their experience with case-based tutorial sessions.

The qualitative analysis generated, as per this study’s conceptual framework: Differentiators of Pharmacology Teaching Modalities (Figure 4), three interlinked themes namely: Variation of experience, Reinforcement learning, and Level of realism. Within the Variation of experience theme, two categories were identified: Interactions and Movement. As for the Reinforcement of learning theme, it included the following categories: Knowledge, Skills, and Attitudes. Lastly, within the Level of realism theme, the following categories were identified: Fidelity and Immersiveness.

**Figure 4.**
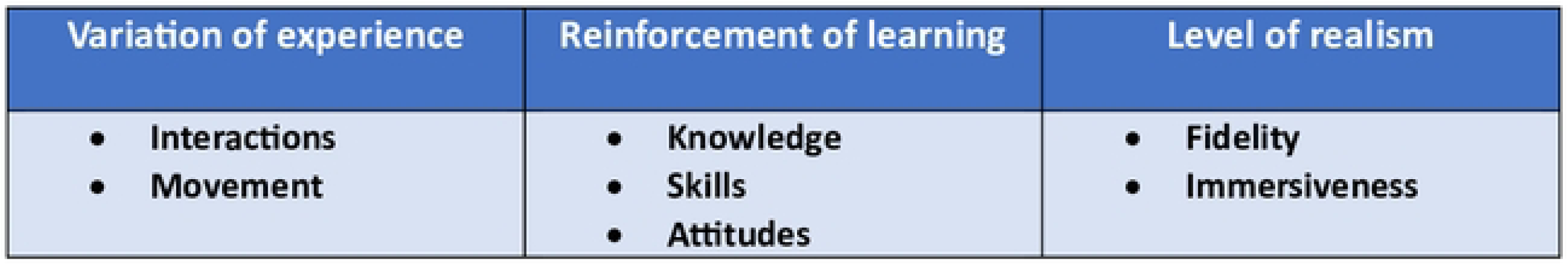
The study’s conceptual framework: Differentiators of Pharmacology Teaching Modalities.

The semi-quantitative tally of text fragments showed the following distribution: Variation of experience (n=8), Reinforcement of learning (n=16), and Level of realism (n=8) (Table 3).

**Table 3.**
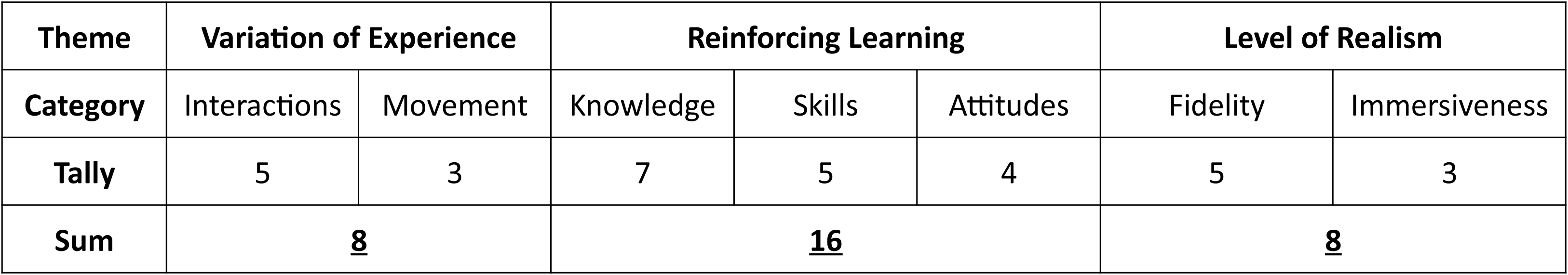
Semi-quantitative tally of the output of the participant-focused qualitative analysis.

| Theme | Variation of Experience |  | Reinforcing Learning |  |  | Level of Realism |  |
| --- | --- | --- | --- | --- | --- | --- | --- |
| Category | Interactions | Movement | Knowledge | Skills | Attitudes | Fidelity | Immersiveness |
| Tally | 5 | 3 | 7 | 5 | 4 | 5 | 3 |
| Sum | <u>8</u> |  | <u>16</u> |  |  | <u>8</u> |  |

#### Variation of experience

This theme encapsulated fragments of text which highlight the students’ perception of the differing encounters and happenings that they experienced through participating in the respective pharmacology course.

##### Interactions

This category included text excerpts that showed students’ reflections regarding the dynamic exchanges and engagements within the session environment.

> 24/S: “…Interacting with patients made me realize the importance of making swift decisions in the clinical environment…”

In terms of the level of interactions, it was clear from the qualitative analysis that the majority of students favoured the high-fidelity simulation over the case-based tutorial sessions.

> 7/S: “…The simulation allowed us to implement what we had learned in class in the form of clinical cases that we are expected to encounter in our future practice; this made the session very interesting and fostered our interactions…”
>
> 84/T: “…I found the simulation more interactive and engaging than the tutorial. I felt that I had immediately integrated the information during the high-fidelity simulation session. The more traditional method of case-based tutorial sessions is not as engaging, and although it involves group work, the learning experience integral to tutorials feels more like a typical lecture…”

Even though most of the students seemed to prefer high-fidelity simulation, there were some students who commended the increased interactions amongst each other and with the faculty members in the case-based tutorial session and reflected on how this enabled self-regulated learning.

> 66/T: “…The intragroup discussions, that were later followed by discussing our answers with the other groups (i.e., intergroup discussions) and with the professors, were very useful and helped me assess my understanding so I know how best to improve my performance…”

##### Movement

This category encompassed the students’ reflections on the physical activity and mobility within their learning environment.

> 22/S: “…We navigated through real-life scenarios, honing our skills in administering drugs. This hands-on experience heightened our awareness of crucial pre-administration details, emphasizing the specific modes of drug delivery and the importance of proper preservation techniques.…”

Relevant to movements, changes, and shifts, some students offered insightful recommendations to enhance the sessions’ effectiveness, proposing ideas to make the learning experience more beneficial for everyone involved.

> 56/T “…Perhaps introducing a multimedia element, such as incorporating pictures or videos for us to analyze, could maximize dynamism in our learning experience, this will add a hands-on dimension to our problem-solving process, fostering a more interactive and immersive educational environment…”
>
> 70/T: “…Imagine if our tutorials adopted a Problem-Based Learning (PBL) or flipped learning style instead of traditional lectures. This would enable student-guided exploration of long cases, analysing them independently and/ or collaboratively. This shift in approach would not only deepen our understanding but also foster a sense of movement as we actively dissect and solve real-life scenarios…”

#### Reinforcement of Learning

This theme revolved around the students’ reflections on the different ways by which their learning experiences were strengthened through the respective teaching modalities. The students alluded to the knowledge gained, skills developed, and attitude shifts that they noticed.

##### Knowledge

This category revolved around students’ thoughts about the acquisition and comprehension of information, facts, and concepts. This category underscored the value that the students associate with deliberate, structured approaches to consolidate knowledge, leading (in the students’ opinion) to more robust and enduring learning outcomes. The students alluded to how the respective modalities affected their capacity to retain knowledge.

> 2/S: “…Despite my oversight in revising, I found the session highly beneficial. I believe that actively engaging with the material in this manner will enhance my retention of the information…”
>
> 10/S: “…Engaging with practical applications of the lesson significantly enhanced my comprehension. I find it more effective to learn by visually observing and actively identifying concepts, rather than by solely relying on classroom problem-solving…”

They also reflected upon how the modality affected their capacity to put into practice the knowledge that they had acquired.

> 51/T: “…The session facilitated a deeper comprehension of both the drugs themselves and the various routes of administration. This practical application of concepts allowed me to grasp the intricacies and nuances involved, leading to a more comprehensive understanding of the subject matter…”
>
> 24/S: “…Interacting with patients provided me with a first-hand understanding of the urgency in making decisions and the rationale behind selecting specific administration routes over others. It also highlighted the practical convenience of selecting certain drug delivery methods…”

##### Skills

This category pertained to the students’ ideas about the practical application and reinforcement of learned concepts. Furthermore, it emphasized the importance of having a holistic approach which integrates theoretical knowledge with practical proficiency to comprehensively attain the learning outcomes.

> 31/S: “…We acquired practical expertise in administering medications within real-world scenarios. Additionally, we delved into pivotal pre-administration considerations, encompassing a comprehensive understanding of the appropriate mode of delivery for each specific drug, as well as the critical aspect of ensuring the optimal preservation conditions for these medications…”
>
> 16/S: “…Engaging with these scenarios proved instrumental in enabling me to apply theoretical knowledge to actual real-life situations. It honed my ability to think swiftly and make critical decisions, particularly in high-stakes, life-threatening situations…”

##### Attitudes

This category encompassed students’ disposition, motivation, and mindset toward the learning that occurred through both modalities. It shed light on the critical role of nurturing a constructive learning attitude for reinforcement and optimization of the learning process.

> 37/S: “…This session offered me a valuable glimpse into the realities of medical practice. It provided a first-hand understanding of the day-to-day operations, decision-making processes, and the intricate dynamics involved in the field. This practical exposure went beyond theoretical knowledge, offering a more vivid and tangible perspective on the intricacies of working in a medical setting…”
>
> 60/T: “…The tutorial cases served to reinforce the main points and objectives covered in the lecture. This experience has significantly broadened my understanding of the inner workings of medical practice, and I am truly grateful for the opportunity it provided. It has illuminated the fact that there is a wealth of knowledge to be gained beyond the confines of textbooks, and I am genuinely enthusiastic about embracing it all. The hands-on exposure has sparked a genuine passion within me for the field, and I am eager to delve even further into the practical applications of this knowledge…”

The students’ attitudes indicated a preference for high-fidelity simulation sessions over case-based tutorial sessions, underscoring their enthusiasm for hands-on, interactive learning experiences.

> 74/T: “… The tutorial cases not only supported the lecture content but also enhanced my understanding. The simulation made the information more accessible and easier to grasp. The interactive nature of the simulation contributed significantly to my learning experience…”
>
> 89/T: “…Unlike a conventional tutorial, the simulation aspect facilitated a hands-on approach, making complex information more digestible and accessible. This not only supported the content from the lectures but also fostered a more comprehensive and practical understanding of the material. Overall, the tutorial cases offered a unique and effective learning opportunity that surpassed the typical tutorial session in terms of interactivity and depth of understanding…”

#### Level of Realism

This theme revolved around the students’ perception of the degree of authenticity and practical relevance incorporated into educational activities. It emphasized the value that the students put on bridging theoretical knowledge with real-world applications to create meaningful and impactful learning experiences.

##### Fidelity

This category denoted the students’ opinions on the learning environment around high-fidelity simulation that mimicked real-world conditions. The students shared their perceptions of the importance of offering them those true-to-life learning experiences, and how they believe it enhanced their educational development. According to the students, the real-life scenarios fed directly into improving their learning experience.

> 4/S: “…I found great satisfaction in translating theoretical concepts into practical applications. Witnessing the application of our classroom learning in a scenario that replicated real-life situations not only motivated my active participation but also stimulated a deeper and more engaged learning experience…”
>
> 9/S: “…It served to fortify crucial concepts and provided us with an opportunity to immerse ourselves in real-life scenarios. This practical exposure is invaluable, equipping us with the skills to intervene effectively and conduct swift analyses when faced with similar conditions in actual patient situations…”

The students also discussed how the real-life scenario and hands-on learning enabled putting the theoretical knowledge into practice.

> 13/T: “…It proved to be an enriching experience, offering me a tangible, real-life outlook on the theoretical knowledge I had acquired. This practical application not only solidified my understanding but also allowed me to connect the dots between classroom learning and its real-world implications, making the educational experience more meaningful and relevant…”
>
> 61/T: “…The session goes beyond mere theoretical instruction by presenting us with potential real-life scenarios to apply the knowledge we’ve gained. What makes this experience particularly valuable is its ability to integrate concepts from various courses, allowing us to grasp the bigger picture…”

##### Immersiveness

This category reflected the student’s learning experience in terms of where the educational environment engaged the senses and created a deeply involving learning atmosphere. It emphasized the importance of creating immersive environments to augment the realism and impact of educational experiences.

> 62/T: “…We have cultivated the mindset of a pharmacist. These sessions have not just been about absorbing facts; they have trained us to think critically. From medication management to dosage calculations, we have learned to analyse medical information with precision, considering factors like drug interactions and patient well-being…”

The students reflected on how engaging in real-life scenarios during the sessions induced stress as they grappled with the challenges that mirror the sophistication of the healthcare environment.

> 23/S: “…This approach proves invaluable in preparing for the challenges of a stressful hospital setting, providing a more realistic and dynamic learning experience. It allows us to seamlessly transition from theory to practice, fostering a deeper understanding and better equipping us to navigate the complexities of our future roles in healthcare…”
>
> 23/S: “…In essence, the stress we encounter in these sessions is constructive, molding us into more resilient and adept future pharmacists. It not only deepens our understanding but also instils a sense of confidence, knowing that we are better equipped to handle the demands of our profession when the stakes are high…”

### Output of quantitative analysis- students’ perception

Two closed-ended questions from the respective survey were utilized to investigate the students’ perspectives of their experience with high-fidelity simulation compared to the case-based tutorial sessions after they completed sessions of both modalities. For the first question about the preferred method of learning pharmacology, the majority of students (90.5%) preferred the high-fidelity simulation method over the traditional case-based tutorial session (9.5%). Responses to the second question revealed that 82.5% of the students found high-fidelity simulation to be more useful, in comparison to case-based tutorial sessions (17.5%) (Figure 5).

**Figure 5.**
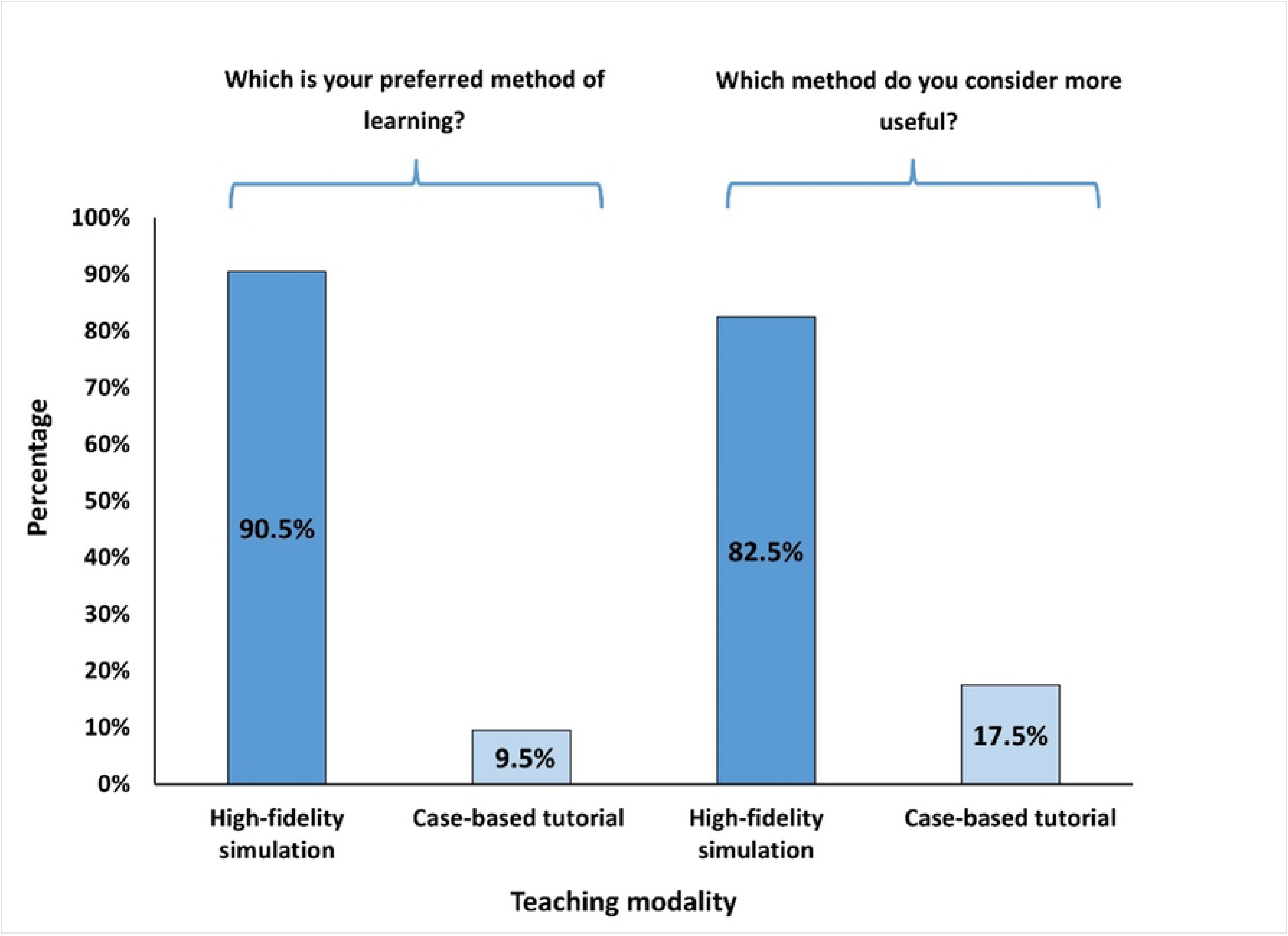
Bar graph illustrating the students’ preference and perceived usefulness of high-fidelity simulation versus case-based tutorial sessions for delivery of pharmacology teaching.

#### Integration

Integrating the outcomes of the thematic analysis with those of the quantitative analyses revealed a comprehensive understanding of the situation, illustrated in the study’s side-by-side joint display (Figure 6). The merging of findings (i.e., primary inferences) enabled the development of a thorough understanding of the students’ experience with high-fidelity simulation and case-based tutorial sessions; it brought together findings about the students’ perception and their performance. As such, the integration led to three groupings of meta-inferences: Satisfaction, Preferences, and Retained Knowledge. The students expressed, in the narrative data, satisfaction with and appreciation of both teaching modalities. There appeared to be no such observations in the output of quantitative analysis. Accordingly, for the ‘Satisfaction’ meta-inference, the integration led to an expansion of the overall viewpoint. For the students’ preferences, the qualitative analysis highlighted that the students favour high-fidelity simulation considering the attributes that they value (namely: variation in the learning experiences, means by which the modalities reinforced the learning, and the extent to which the learning environment mimicked reality). Along those lines, part of the quantitative data analysis showed that the students prefer high-fidelity simulation and find them more useful. This shows that for the ‘Preferences’ meta-inference, the integration allowed for both confirmation and expansion of the overall viewpoint. As for the effect of the teaching modality on the retention of knowledge, the quantitative analysis showed that there is no statistically significant change in knowledge retention between both modalities, whilst the qualitative data analysis showed that the students perceive the high-fidelity simulation to reinforce learning more than case-based tutorial sessions. Accordingly, for the ‘Retained Knowledge’ meta-inference, the integration led to a refinement in the overall viewpoint.

**Figure 6.**
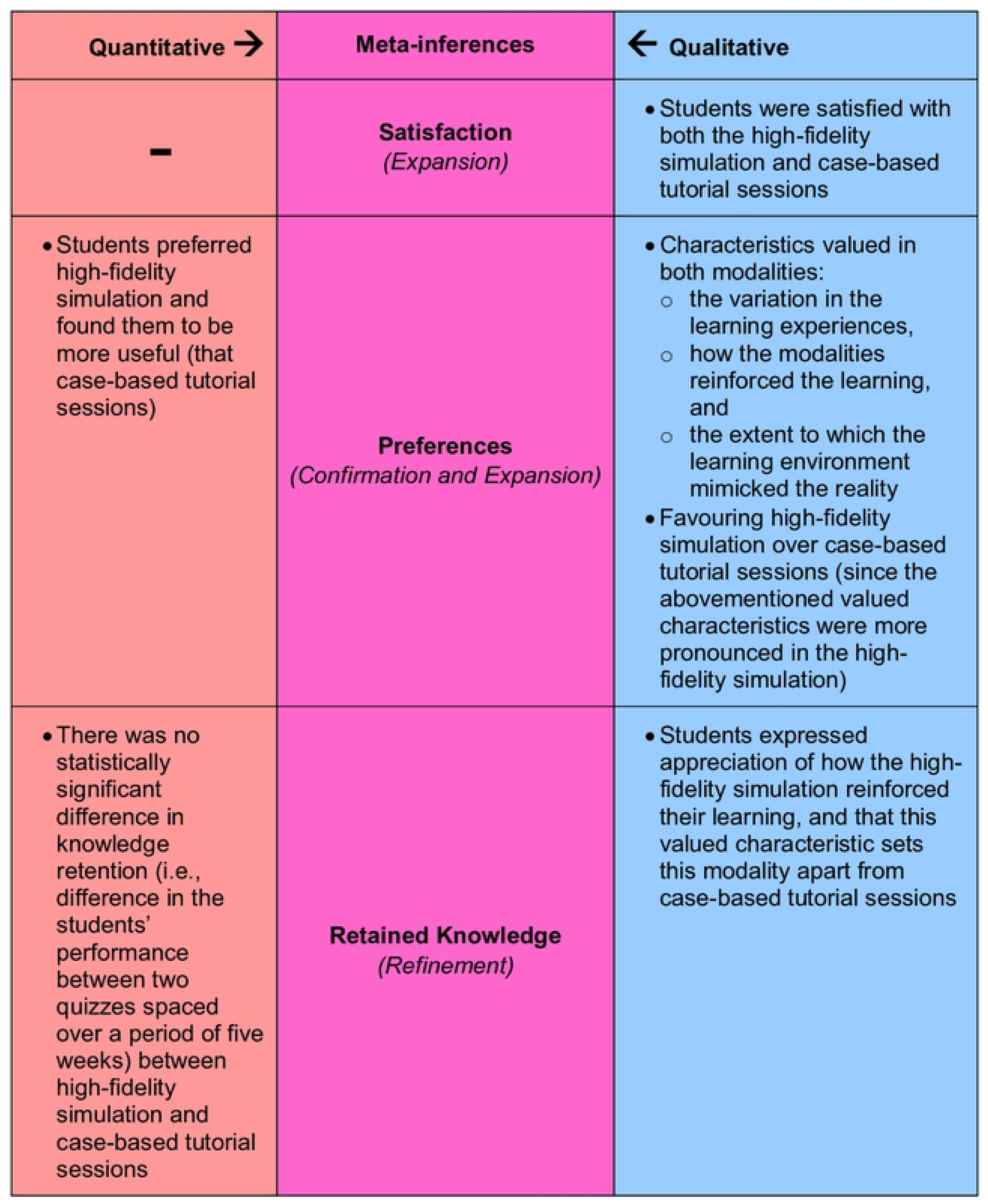
Output of the iterative joint display analysis process, resulting in three meta-inferences: Satisfaction, Preferences, and Retained Knowledge. The secondary colour Purple emerged by mixing the primary colour Red with the primary colour Blue (which constitutes an analogy of the lateral and critical thinking that took place to generate the meta-inferences from the integration of two sets of primary inferences). For the ‘Satisfaction’ meta-inference, integrating the quantitative with the qualitative inferences led to an expansion in the overall viewpoint. For the ‘Preferences’ meta-inference, the integration led to both: confirmation and expansion of the overall viewpoint. As for the ‘Retained Knowledge’ meta-inference, the integration led to the refinement of the overall viewpoint.

## Discussion

This study adds to the paucity of literature regarding the incorporation of high-fidelity simulation into the pharmacology course in undergraduate medical studies. Most studies regarding the effects of high-fidelity simulation in comparison to case-based tutorial sessions rely on observations, without any empirical research component (40). The current study investigated the impact of high-fidelity simulation and case-based tutorial sessions on Year 2 medical students’ retention of pharmacology knowledge. Additionally, the study explored the personal experience of individual students and their subjective perception of both teaching methods. The results of this study revealed no statistically significant difference in knowledge retention, as measured by performance on the 10-MCQ quiz, between the two teaching modalities. The participating undergraduate medical students were satisfied with both high-fidelity simulation and case-based tutorial sessions for pharmacology teaching, while favouring the high-fidelity simulation modality and finding it to be more useful. Students brought up three groups of characteristics that they valued in both the high-fidelity simulation and case-based tutorial sessions. These attributes include: the variation in the learning experiences that both modalities offer, how the respective modalities reinforce the learning, and the extent to which the learning environment (characteristic of each modality) mimics the reality. Collectively, those attributes, constitute a novel conceptual framework, namely: Differentiators of Pharmacology Teaching Modalities, that the current study introduced. This framework can enable health professions’ educators in systematically considering, when designing pharmacology curricula, what students perceive to differentiate teaching modalities.

In alignment with previously conducted studies (41–43), the quantitative analysis of the current study showed that the choice of modality did not affect the students’ knowledge retention. However, the qualitative analysis revealed that the students perceived the high-fidelity simulation to reinforce their learning of pharmacology more than case-based tutorial sessions. Similar findings have been observed in a previously conducted randomized controlled trial which compared simulation-based learning to traditional lectures in teaching the diagnosis and management of bronchial asthma (43). Thus, it is worth considering both the modality’s effect on students’ performance and the students’ perception of the learning experiences in deciding whether or not to adapt it.

For the students participating in this study, both modalities introduced variations in the learning experiences in comparison to traditional lectures, yet the high-fidelity simulation was perceived to be more interactive, entailing more physical movement. Along those lines, a previously conducted mixed methods cross-over study showed that students appreciated the movement and interactions integral to high-fidelity simulation (44). Moreover, another study which relied on the feedback of 103 fifth year medical students, who took part in a simulation training in nephrology, revealed that the students’ appreciated the dynamism integral to high-fidelity simulation and found it to be interesting (45). Relevantly, a prospective crossover observational study showed that high-fidelity simulation was superior (relative to other teaching modalities) in harvesting leadership, teamwork, and task management skills (46). Also, a thematic analysis of data collected from interviews with students identified three types of student engagement in the simulation-based learning environment: reflective engagement, performance engagement, and interactive engagement (47).

Although, in the current study, both modalities appeared to contribute to reinforcing competencies, including knowledge, skills, and attitudes, high-fidelity simulation was perceived by the students to be more effective in this regard. Similarly, a previous examination of 177 undergraduate nursing students in recognizing and responding to hypovolemia showed that high-fidelity simulation develop various competencies including knowledge about the subject matter and attitudes (e.g., self-confidence) (48). Another study also proved the value of high-fidelity simulation on the development of skills, including but not limited to flexible and reflective thinking (49).

The current study also showed that both modalities were thought to bring the subject matter closer to reality, although the high-fidelity simulation was considered significantly more immersive. It is worth reiterating that simulation has been increasingly used by medical schools to provide students with a safe environment to practice various skills, ranging from hands-on medication administration to complex decision-making skills that mimic real-life clinical scenarios (50). Simulation comes in various levels of fidelity. Low-fidelity simulation encompasses basic mannequins or computer programs that represent simplified aspects of real-world scenarios. Medium-fidelity simulation includes more advanced models and virtual environments that offer a closer approximation to real-life scenarios. Finally, high-fidelity simulation (as previously described in the Introduction section) utilizes advanced, often interactive technology and realistic human mannequin to closely mimic real-world scenarios, providing an immersive learning experience. Yet, there remains a dilemma concerning the effectiveness of high-fidelity simulation when compared to case-based tutorial sessions in pharmacology education, especially when considering cost-to-benefit ratio, given that high-fidelity simulation is costly (51), and requires ample of time and resources in both the planning and implementation phases.

It is suggested that appraising the value of simulation-based medical education requires complete accounting and reporting of cost. Various methods of cost analysis exist, each meant to address a focused economic research question (24). A previously conducted systematic literature review, aimed at summarizing studies that contain an economic analysis of simulation-based medical education for training of health professions learners, showed that most studies focused on the equipment and materials cost, mainly the price of the simulator, and hence, were of the most limited form of cost analysis: basic cost/cost feasibility analysis (52). It is suggested that this cost analysis (i.e., evaluating only cost) is useful if the evaluator wants to simply know how much a particular learning and teaching intervention costs, and whether it can be executed within budgetary constraints. However, if the evaluator wants to be able to reach conclusions not solely about cost, but also about the relative benefits or effectiveness of a range of interventions, a cost-benefit (i.e., evaluating costs and monetary outcomes) or cost-effectiveness (i.e., evaluating costs and educational outcomes) is required (53). From this perspective, it is important to consider leveraging high-fidelity simulation only where it is likely to add sufficient value (in terms of benefit, effectiveness, and utility), which requires a full-economic evaluation (54). For instance, high-fidelity simulation has been shown to improve decision-making skills, as well as support in learning effective communication with patients, and thus its use in such instances is greatly encouraged (55). Moreover, there is a subset of medical education where simulation-based learning has demonstrated great superiority over traditional lecture-based sessions. Such examples include more hands-on skills such as surgical specialties, obstetrics emergencies, and the use of medical equipment such as ultrasound (56, 57).

The students’ reflective descriptions of their experiences with high-fidelity simulation in pharmacology teaching reveal the value of designing the learning intervention in alignment with both Kolb’s experiential learning cycle (13, 14) and social constructionism learning theory (18, 19). Relying on those experiential learning theories enabled maximizing the reflections (and in turn development) that the students go through on an individual basis as part of the experience, along with the growth that occurs due to the students’ active participation and engagement with others while immersed in the environment of learning.

The current study exhibited the value of deploying mixed methods research in evaluating the efficacy and effectiveness of learning and teaching interventions. This research design enabled tapping into more than one level of analysis which offered a systemic perspective of the students’ learning experience, better informing associated decisions and continuous quality enhancement activities (58). This is in alignment with the literature that encourages employing models such as Kirkpatrick (31) and LTEM (32) when deciding on mechanisms of evaluating learning and teaching interventions (which ideally needs to be effectively done as part of the planning for the respective learning and teaching interventions). In conjunction with mixed methods research design, such evaluation models hold ample of potential in raising the rigor of institutional research activities conducted primarily for performance improvement in medical education (58). Besides the various levels of analysis that were captured via the chosen research design, the output of the integration covered more than one level, as well, where ‘satisfaction’ and ‘preferences’ are more related to the students’ reaction to the learning experience, and ‘retained knowledge’ corresponds mostly to the competencies developed as a consequence to the learning and teaching intervention.

This study showed that both modalities fostered self-regulated learning (10, 59), where (according to the students) high-fidelity simulation was more influential (relative to case-based tutorial sessions) in this regard. The students expressed a lot of enthusiasm around the interactive, hands-on learning experiences integral to high-fidelity simulation, that were designed to catalyse students’ reflective practice and their active participation in the experiences themselves.

This study has a few limitations. Although the choice of mixed methods research design generated in-depth insights, the generalizability of the findings is limited to universities that are contextually similar to MBRU. Hence, it is recommended for future studies to include multiple medical colleges, and/or several cohorts within the same college. Such a study can be longitudinal in nature to allow for investigating causality between variables since this study was restricted to uncovering associations. Lastly, the current study mainly focused on knowledge retention assessed by multiple-choice questions, leaving room for future research to explore other dimensions of learning outcomes, both specific and non-specific to pharmacology courses including practical skills application and clinical decision-making.

## Conclusion

This research investigated the impact of high-fidelity simulation and case-based tutorial sessions on knowledge retention of Year 2 medical students in pharmacology, as well as their perception on both modalities. While there was no statistically significant difference in students’ knowledge retention between high-fidelity simulation and case-based tutorial sessions, the study highlighted students’ preference for high-fidelity simulation, given the variation of experiences, reinforcing of learning, and extent of realism that they believe was offered by high-fidelity simulation compared to case-based tutorial sessions. This brings forth the value of anchoring the design of high-fidelity simulation in experiential learning theories. The study advocates caution in adapting high-fidelity simulation, where careful evaluation can lend itself to identifying contexts where it is most effective.

## Data Availability

All relevant data are within the manuscript and its Supporting Information files.

## Disclosure

The authors report no conflicts of interest in this work.

## Acknowledgements

The authors would like to thank the Year 2 medical students who participated in this research, and to acknowledge the valuable statistics’ advice that they received from Professor Jeyaseelan Lakshmanan. They would also like to extend gratitude to Professor Glenn Matfin and Ms. Helen Henderson, along with the rest of the staff members at the Khalaf Ahmad Al Habtoor Medical Simulation Center at MBRU for their support in the planning and implementation of the learning and teaching interventions reported upon in the current study.

## Notes

### Competing Interest Statement

The authors have declared no competing interest.

### Funding Statement

The author(s) received no specific funding for this work.

### Author Declarations

Ethical approval for the current study was granted by the Mohammed Bin Rashid University of Medicine and Health Sciences-Institutional Review Board (MBRU-IRB-2020-001), and electronic informed consent was obtained from all participants prior to participating in the research study.

